# Barriers and enablers to improving integrated primary healthcare for non-communicable diseases and mental health conditions in Ethiopia: a mixed methods study

**DOI:** 10.1101/2023.11.21.23298770

**Authors:** Alemayehu Bekele, Atalay Alem, Nadine Seward, Tigist Eshetu, Tewodros Haile Gebremariam, Yeneneh Getachew, Wondosen Mengiste, Girmay Medhin, Lara Fairall, Nick Sevdalis, Martin Prince, Abebaw Fekadu, Charlotte Hanlon

## Abstract

**Background:** The Ethiopian Primary Healthcare Clinical Guidelines (EPHCG) seek to improve quality for people with Non-Communicable Diseases and Mental Health Conditions (NCDs-MHCs) and provide an integrated approach to multi-morbidity. The aim of this study was to identify barriers and enablers to implementation of the EPHCG with a particular focus on NCDs-MHCs.

**Methods:** A mixed-methods convergent-parallel design was employed from May, 2019 to January, 2020 after implementation of EPHCG in 18 health facilities across four districts and one town administration in southern Ethiopia. Semi-structured interviews were conducted with 10 primary healthcare clinicians and one healthcare administrator to identify barriers and enablers. The Organisational Readiness for Implementing Change (ORIC) questionnaire was self-completed by 124 health workers to identify facility level readiness for change. Determinants from both approaches were mapped to the Consolidated Framework for Implementation Science (CFIR) and the Theoretical Domains Framework (TDF). Expert Recommendations for Implementing Change (ERIC) were employed to select potential implementation strategies to address barriers.

**Results:** Four thematic domains, EPHCG training and implementation, awareness and meeting patient needs (demand side), resource constraints/barriers (supply side) and care pathway bottlenecks were identified. The innovative facility-based training to implement the guidelines had a mixed response, especially in busy facilities where teams reported struggling to find protected time to meet. Key barriers to implementation of EPHCG were non-availability of resources (CFIR inner setting), such as reagents for laboratory tests and medications that undermined efforts to follow guideline-based care; the way care was structured and lack of familiarity with providing care for people with NCDs-MHCs. Substantial barriers arose because of interlinked socio-economic problems that were interlined with health but not addressable within the health system (CFIR outer setting). Several behavioural determinants influenced effective implementation of EPHCG (TDF), including low population awareness about NCDs/MHCs and unaffordable diagnostic and treatment services. Implementation strategies were identified. Survey findings indicated high scores of organisational readiness to implement the desired change but were notably more positive than the qualitative data.

**Conclusions:** Although perceived as important and necessary, practical implementation of EPHCG was constrained by challenges across domains of internal/external context and behavioural determinants. This was especially marked in relation to expansion of care responsibilities to include NCDs-MHCs. Attention to social determinants of health outcomes, community engagement and awareness-raising are needed to maximize population impact.

## Introduction

Primary healthcare (PHC) is at the heart of efforts to achieve Universal Health Coverage and has three principal pillars: community empowerment, multisectoral policies and action, and integrated delivery of quality primary care and public health services to people in need (1). For comprehensive realisation of this vision, it is vital for PHC to address the ever-increasing population burden of chronic conditions in low- and middle-income countries (LMICs), especially non-communicable diseases (NCDs) and mental health conditions (MHCs). The burden of NCDs is increasing substantially in Ethiopia, accounting for 39.3% of all deaths and 34% of Disability-Adjusted Life Years (DALYs) in 2016 (2). DALYs due to MHCs were estimated at 2.2% per the projection made by the WHO for Ethiopia (3). Reliance on vertical programmes or centralised models of specialist care has resulted in high treatment gaps for both NCDs (4) and MHCs (5) and a fragmented and inadequate response to co-morbidity or multi-morbidity (6). In response, it has been recommended to integrate care for people with NCDs and MHCs into service packages delivered within PHC (7,8).

Presently, the Ethiopian Ministry of Health (MoH) is planning to expand access to care for people with NCDs/MHCs through integration in PHC (9). The high co-morbidity and multi-morbidity within and between NCDs/MHCs means that integrated care promises better healthcare outcomes (10,11). In the Chronic Care Model (12), adapted for LMICs as the innovative care for chronic conditions framework (ICCCF)(13), co-ordination of care to respond to the individual’s multiple needs and preferences (person-centred care), empowerment of patients to be active partners in care, and systems that support treatment-to-target are associated with better health outcomes. In recognition of this, the Ethiopian MoH localised the South African Practical Approach to Care Kit (PACK) programme to become the Ethiopian Primary Health Care Clinical Guidelines (EPHCG) (14). EPHCG is a set of evidence-aligned guidelines that are designed not just to standardise the care provided at PHC level but also to horizontally integrate care so that PHC workers are prompted to consider co-morbid and multimorbid conditions at every visit. EPHCG furthermore provides guidance for the longitudinal assessment and treatment of care needs. Other innovative implementation approaches of the guidelines include facility-based training involving the whole PHC team using adult learning principles, case based (practical application) and reflection (14).

Following localisation of the EPHCG(14), training was cascaded through master trainers and facility trainers who then convened facility-based, peer-led training in over 550 primary care health centers in Ethiopia over a 12-month period. Initial programmatic evaluation indicated that EPHCG was well-received but that impact was undermined by weak leadership at district and health facilities level and lack of PHC workers commitment, shortage of resources, high caseloads, and misperceptions about onsite training (15). Strengths of this implementation included the strong involvement of top leadership, fair budget allocation for implementation, and integration within the existing health system. However, implementation barriers and enablers, including organisational readiness, were not evaluated systematically.

Organisational readiness to change is recognised as critical to successful implementation of new initiatives. Organisational readiness is a multi-level, multi-faceted concept referring to a shared commitment of members of an organisation to implement a change and a shared belief in their collective capability to do so (16). Readiness varies as a function of how much organizational members value the change and how favorably they appraise three key determinants of implementation capability: task demands, resource availability, and situational factors (17). As well as overlooking readiness, previous investigations of EPHCG implementation did not examine the specific impact on care for people with NCDs/MHCs.

The aim of this study, therefore, was to identify barriers and enablers experienced in the implementation of the EPHCG with particular focus on care for NCDs/MHCs.

## Methods

### Study design

A mixed methods convergent parallel design (18) was employed. Qualitative data were collected from EPHCG training participants using key informant interviews of. Quantitative data were obtained from a cross-sectional assessment of organisational readiness administered to all PHC workers in the selected health facilities after completing facility based EPHCG training. The quantitative and qualitative components were conducted contemporaneously. Findings of the two components were analysed independently but integrated through mapping onto two implementation research determinant frameworks: the Consolidated Framework for Implementation Research (CFIR) (19) and the Theoretical Domains Framework (TDF) (20). Implementation strategies for the determinants mapped were also recommended using the Expert Recommendations for Implementing change (ERIC) tool (21).

### Study period

The EPHCG training at facility level was conducted in 2018 and continued up to April 2019 in the study setting. The training in each health facility lasted eight weeks i.e., 1-2 hours per week for eight weeks. The training was facility based (onsite), participatory, case based and used adult learning principles (22). The data collection for both qualitative and quantitative assessments were conducted from May 2019 to January 2020.

### Study setting

The study was conducted as part of the Health System Strengthening in sub-Saharan Africa (ASSET) programme(23).The setting was four districts and one town administration of the Gurage Zone, Southern Nations, Nationalities and People’s Region of Ethiopia: Meskan, Misrak Meskan, Sodo and South Sodo districts and Butajira town. These districts are located around 100 to 130km south of the capital city, Addis Ababa, and are predominantly rural (24). The total population size of these districts is estimated to be 404,809 in 2017(25). Health services are provided by one general hospital (Butajira hospital), one primary hospital (Buee hospital), 16 health centres and 94 health posts (26). PHC services are rendered through the PHC Units, comprising a health centre and 3-5 linked health posts. Health centres are staffed by an average of 20 health personnel, including nurses and health officers, and provide both preventive and curative services that focus mostly on infectious diseases, maternal and child health conditions and nutritional problems. Health posts are closer to the community and focus on health promotion and illness prevention. They are staffed by two female health extension workers with 1-2 years of basic training. Community based health insurance had been recently initiated at the time of this study. The intention to integrate care for people with MHCs and NCDs into primary health care services has been articulated in national level strategies from the Ministry of Health (7).

### (1) Qualitative study

#### Sampling

We sampled healthcare workers based in health centres where EPHCG training was conducted, purposively selecting respondents in terms of rural versus urban health centres and their professional role within the facility. We additionally sampled healthcare administrators involved in co-ordination of the EPHCG implementation.

#### Eligibility criteria

Health workers and healthcare managers who completed the EPHCG training and who had been tasked with implementing the guidelines were included.

#### Data collection and instruments

Semi-structured interviews were conducted using an interview guide that focused on training and use of EPHCG, perceived needs of people with NCDs/MHCs, care pathways and information systems for people with NCDs/MHCs, and possible health system strengthening interventions to address bottlenecks. EPHCG review meeting minutes and field notes were also recorded and used as a data source. Notes and audio-recording were used to capture the interview discussions.

#### Data analysis

For the qualitative study, interviews and meeting minutes were transcribed in Amharic, translated into English and uploaded to Open Code 4.03 software (27). Thematic analysis was carried out by understanding the data, generating the initial codes, searching for themes, reviewing the themes, defining the themes and producing the results. The data from thematic analyses were then mapped to the theoretical domains. Contextual determinants were mapped to the Consolidated Framework for Implementation Science (CFIR) (19) and potential determinants of behaviours were mapped to the Theoretical Domains Framework (TDF) (20). Potentially relevant implementation strategies were selected using findings from the Expert Recommendations for Implementing Change (ERIC) project (28). ERIC provides a taxonomy of implementation strategies that were developed using a Delphi process.

### Cross-sectional study of organisational readiness

#### Study population

For the cross-sectional study, we approached all healthcare workers in facilities where EPHCG implementation was underway and district health administration focal persons for the EPHCG implementation.

#### Sampling and sample size

The target sample size of 200 was based on the estimated total number of health workers 18 health facilities across four districts and one town administration in southern Ethiopia. **Data collection and instruments**

The Organisational Readiness of Change (ORIC) questionnaire was used (29). ORIC seeks to identify barriers/enablers (i.e., determinants) to implementation. It is a self-report questionnaire comprising 12 items: 7 items related to ‘change efficacy’ and 5 items related to change commitment. Change efficacy refers to the shared belief of members of the organisation in their “collective capabilities to organise and execute the courses of action involved in change implementation” (30). This concept reflects the amount of knowledge available about what to do and how to do it; i.e., it is a function of members’ cognitive appraisal of three aspects of implementation capability: the resources (including time) available, task demands, and the situation the organisation faces. The ability to measure how an organisation’s members perceive these two constructs may, ultimately, be used by health care leaders to develop more effective and efficient change strategies (29). Change commitment can be defined as the mindset that binds an individual to the course of action deemed necessary for the successful implementation of a change initiative (31). Commitment was also described as organisational members’ willingness and motivation to attain the change they need to ultimately solve problems and promote the desired behaviours for change (17). We provided the questionnaires in envelopes with a unique individual identifier and a code to indicate the health facility. Respondents were invited to complete the survey independently and return to the data collector in a sealed envelope.

### Data analysis

Descriptive analyses of sociodemographic characteristics and variables were computed. The total scores for the change-efficacy and change-commitment subscales were not normally distributed and the sample size of the study was small. Thus, Kruskal Wallis ranked sum test (32) was used to investigate statistical significance of the median score differences across different factors.

## Results

### Sociodemographic characteristics

A total of 11 study participants whose age ranged from 23 to 35 years participated in the qualitative study: seven males and four females. Six of the 11 study participants were health officers and the remaining were nurses, non-specialist doctors and healthcare administrator (Table 1).

**Table 1:** Sociodemographic characteristics of participants for qualitative study (N=11)

| Characteristics | Response category | Number |
| --- | --- | --- |
| Age (years) | 21-25 | 2 |
|  | 26-30 | 7 |
|  | 31-35 | 2 |
| Gender | Male | 7 |
|  | Female | 4 |
| Professional category | Health Officer | 6 |
|  | Nurse | 3 |
|  | General practitioner | 1 |
|  | Healthcare administrator | 1 |

**Table 2:**
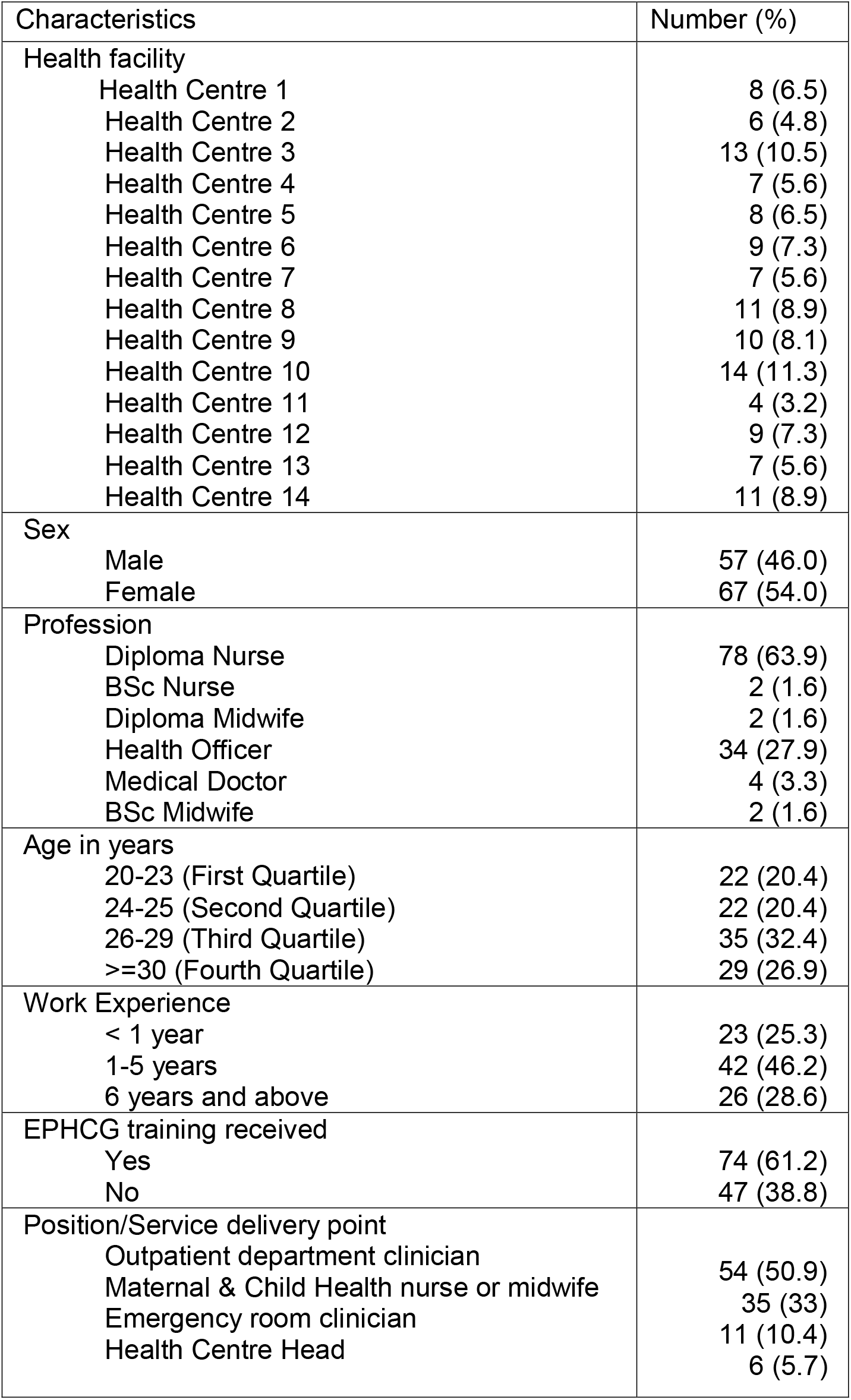
Sociodemographic characteristics of participants in cross-sectional study (n=124)

| Characteristics | Number (%) |
| --- | --- |
| Health facility |  |
| Health Centre 1 | 8 (6.5) |
| Health Centre 2 | 6 (4.8) |
| Health Centre 3 | 13 (10.5) |
| Health Centre 4 | 7 (5.6) |
| Health Centre 5 | 8 (6.5) |
| Health Centre 6 | 9 (7.3) |
| Health Centre 7 | 7 (5.6) |
| Health Centre 8 | 11 (8.9) |
| Health Centre 9 | 10 (8.1) |
| Health Centre 10 | 14 (11.3) |
| Health Centre 11 | 4 (3.2) |
| Health Centre 12 | 9 (7.3) |
| Health Centre 13 | 7 (5.6) |
| Health Centre 14 | 11 (8.9) |
| Sex |  |
| Male | 57 (46.0) |
| Female | 67 (54.0) |
| Profession |  |
| Diploma Nurse | 78 (63.9) |
| BSc Nurse | 2 (1.6) |
| Diploma Midwife | 2 (1.6) |
| Health Officer | 34 (27.9) |
| Medical Doctor | 4 (3.3) |
| BSc Midwife | 2 (1.6) |
| Age in years |  |
| 20-23 (First Quartile) | 22 (20.4) |
| 24-25 (Second Quartile) | 22 (20.4) |
| 26-29 (Third Quartile) | 35 (32.4) |
| >=30 (Fourth Quartile) | 29 (26.9) |
| Work Experience |  |
| < 1 year | 23 (25.3) |
| 1-5 years | 42 (46.2) |
| 6 years and above | 26 (28.6) |
| EPHCG training received |  |
| Yes | 74 (61.2) |
| No | 47 (38.8) |
| Position/Service delivery point |  |
| Outpatient department clinician | 54 (50.9) |
| Maternal & Child Health nurse or midwife | 35 (33) |
| Emergency room clinician | 11 (10.4) |
| Health Centre Head | 6 (5.7) |

### Qualitative findings

The findings were grouped into four thematic areas: EPHCG training and implementation, Awareness and meeting patient’s needs (demand side), Resource constraints/barriers (supply side), and Care pathway bottlenecks.

### EPHCG training and implementation

The EPHCG was reported to have a range of benefits. For instance, the presentation of patients with NCDs and/or MHCs reportedly increased after the introduction of the guidelines. However, the confidence of health workers in identifying NCDs and MHCs was variable. Healthcare workers reported that they could easily identify and manage hypertension and asthma but did not feel confident to identify and manage depression, cardiac problems and diabetes.

> *“I know about depression from my school education. There is one course on psychiatry. What I know about depression is what I have got from that course. There is no training here on depression experience or something. I did not take training on depression as a professional.”*

> Participant with ID NCD-03

The quality of care and confidence of healthcare workers was nonetheless reported to have improved after EPHCG training.

> *“…It was great. The quality of the work that is being done in that area is improving. When we compare how we gave treatments before the training versus how we give the treatments now, the result is satisfactory.”*

> Participant with ID NCD-08

> *“I have better confidence, but the main thing is the standard. If the standard orders you to give the treatment here or to refer it, what you do will be based on that. If you sometimes miss some things, it will give you better confidence in reading the guideline.”*

> Participant with ID NCD-07

The pros and cons of the onsite training approach were raised by participants. One perceived advantage was the greater opportunity for participation when compared to the usual “hotel-based training”.

> *“The training provided at other places, hotel or at woreda, and training provided here are different. You may not be heard or get chance to ask questions on training that you will get in other places, or you may not ask what you don’t understand as there will be many participants of the training. But here we will repeatedly discuss until everyone understand it. To me, other than the payment [per diem for off-site training], training provided here is better.”*

> Participant with ID NCD-10

However, there were reflections on the challenges of onsite training. These included a perceived lack of commitment at the health centre level, lack of familiarity with peer learning, difficulty in finding time for training when engaged in their routine work, lack of the financial incentives that accompany off-site training, concerns about the fidelity/quality of peer-led training, and a high turnover of trained trainers and facilitators. Some participants reported being distracted by other tasks:

> *“… it is better to train somewhere [else] to fully catch up with the book. Because here you are on the job and you think of the job. For instance, if you are training in the morning you have to go to work in the afternoon, you are at work. In addition, you may not see what you have trained today. You are at work; you may get tired and sleep. But if you go there, you have taken the training, and everything is about it, nothing else will be there.”*

> Participant with ID NCD-12

> *“There are also health centers that we cannot initiate the training despite our frequent support. We have to support those health centers and take this issue as a challenge. The reasons are the health professionals working in that health center have thought we will not train unless we get incentives as we work in shift. But those health professionals who understand the aim are attending the training.”*

> Participant with ID NCD-14

EPHCG implementation was observed and reported to be variable across districts and health centers, partly related to the extent to which PHC workers understood the purpose of EPHCG. Implementation challenges included not feeling comfortable using the guideline during consultation, inadequate support and supervision and lack of awareness about EPHCG across all levels of health systems. Nonetheless, there were health centers which had made EPHCG an integral part of their work, reinforced by meetings and discussions between the health workers.

> *“Everyone is performing based on the training they took… we see [patient medical] cards and discuss the cases to check whether they are managing cases based on the discussion we had or not…umm. We discuss cases during the morning session and talk about ways on improving them. We are working hard to keep the flow of the EPHCG because everyone is applying what they got from the training…”*

> Participant with ID NCD-17

### Awareness and patient’s needs (demand side)

Successful implementation of the EPHCG for people with NCDs/MHCs was reported to be affected by low patient and community awareness. Preference for traditional or religious healers and low awareness about the availability of the new services in the health centers contributed to late presentations i.e., after complications manifested. Emotional needs were perceived not to be addressed well. There were also reports that raising expectations of improved quality of PHC led to disappointment from the community due to the poor availability of diagnostic tests and medications.

> *“It might look like it is an easy thing but the main problem in the community right now is not having enough awareness. Lack of awareness makes them use local or traditional solutions for problems that could be treated easily with medicine. Maybe people will come here after trying traditional medicines for depression and psychotic disorders. Maybe they will come here to be treated scientifically after the traditional solution failed to do so and education about NCDs should be given to the community because you can make a difference on something if you first work on the behavioral change of the people. If the way of thinking of the community is not changed, there will be no difference and the goal could not be met even if you offer the drug because it should be taken properly.”*

> Participant with ID NCD-17

Low awareness of patients, particularly those from rural settings, was found to be a barrier to implementing person-centered care.

> *“Most of the patients using this health facility are from rural settings, they may not understand what we are talking to them. But even if they are rural people, they could say they prefer injection while they have to take tablets. Then I would explain that the tablet is more important for them instead of the injection and that they could come back if they don’t have any progress and to take other medication. I explain and convince them and let them take the medication.”*

> Participant with ID NCD-09

Respondents reported that there were many people with chronic diseases in the community who could not afford to access the health service. The community-based insurance system was perceived as the good initiative to address this gap.

> *“Chronic illnesses might not be cured shortly. In relation to the cost, it is now better as all of them are using health insurance, they are treated for free. There are people who are considered as very poor and cannot pay 200 birr every one or two years…..”*

> Participant with ID NCD-03

### Resource constraints (supply side)

The supply side of the health service delivery was constrained by many factors. These included the high patient load, shortage of outpatient department (OPD) space, poor availability of laboratory tests for diagnosis, weak systems for monitoring the patients’ conditions and unreliable medication supplies.

> *“There are a lot of patients here and in addition to the shortage of OPDs [out-patient department rooms]. There is also a shortage of blood pressure apparatus and that is a problem. The laboratories do not have reagents for diagnostic tests. There is also a shortage of drugs even if they are supplied every month. Sometimes, some medicines for hypertension and diabetes might not be available. These are some of the problems here…”*

> Participant with ID, NCD-17

The limited number of health professionals to provide the service was another major bottleneck hampering the provision of health services.

> *“…there is a shortage of professionals at this time compared to the high patient flow in the health centre I am working in.… the health centre is operating like a hospital. There are lots of patients but limited number of professionals to provide the required service. As there is also shortage of budget, we face difficulty to be well equipped with the necessary medications.”*

> Participant with ID, NCD-08

The other important dimension was lack of space, specifically to provide care for patients with mental health conditions.

> *“We could get patient with psychiatric problem; we don’t have specific place for them. Instead of talking to them in front of other people, it would have been good if we have a specific place or room for them to come on their follow up date and discuss with us freely without being afraid of other people. The place could also help us provide different entertaining things and better counseling service. So, I recommend if there is separate room for these patients. If the case of the patient is depression, you could take them to other room so that s/he could properly talk more about his/her situation. Sometimes they talk low, sometimes they talk loudly or slowly. So, in order for them to talk freely, it would have been good to have separate room for such cases”*

> Participant with ID, NCD-09

### Care pathway bottlenecks

Within the care pathway, detection of depression was not perceived to be a problem by one of the respondents, although other health workers lacked confidence to make the diagnosis.

> *“…I am mental health focal person to this level and I am working on depression. As I said before depressed patients come with headache or other symptoms. Depression is identified by asking about their symptoms and history in detail. When once diagnosed we do not proceed directly to drug treatment. Initially we start with advice and will give appointment. If they do not improve, we will start drug treatment.”*

> Participant with ID, NCD-15

Participants also reported that some elements of self-management and lifestyle changes needed for people with NCDs/MHCs were happening, but that requires the health worker to engage with family support and to develop an ongoing relationship with the patient.

> *“…. And if the patient is old, we ask their caregiver or whoever brings them to us if the patient is taking the medication properly or not. In addition, we also ask their family, if the patient is properly taking the medication, if he is eating salt or not and if he/she is having physical exercise. Since their families could not clearly tell us the right thing, we try to ask them like a family. Because, since these patients come for follow up every month every two weeks or every week, if we treat them as family, they could open up about everything. We create that family vibe as much as possible and ask their family member. Then they will be free to talk to us and they do not hide any information from us.”*

> Participant with ID, NCD-06

Monitoring and support for adherence to medication among patients diagnosed to have NCDs or MHCs was found to be problematic.

> *“Yes, we don’t usually monitor hypertension and mental health in terms of whether they are taking their pills or not. We do monitor for tuberculosis patients but not for patients with other conditions.”*

> Participant with NCD-09

Concerns about patients being inadequately engaged with care were also expressed.

> *“You will know if they are properly taking their medicines when the progress is good on the adherence. Some patients might come to their follow up a little later, after three or four days skipping their medicine also because they run out of it. We will refer their adherence to this, there are health extension workers in the community.”*

> Participant ID with NCD-07

Follow up of patients with chronic conditions was being monitored through a paper-based registry. This was the mechanism through which health workers could see if people dropped out of care and was workable due to the modest number of people enrolled.

> *“We have registration book. We register patient’s next appointment on both the book and paper that is given to the patients. Therefore, we check by looking at the registration. We even know each patient.”*

> Participant with ID, NCD-06

The role of the health extension workers and the importance of tracing people who dropped out of care was emphasized.

> *“We specially follow the mental [health] cases. We know if the diabetic patients came or didn’t. There are sources of information from the district, they have files of the deceased where causes of death are written. Health extension workers are the ones that follow up the cases in clinics. However, when it comes to quitting of taking medications, besides the mentally ill cases, like I said before, you wouldn’t know unless you trace them.”*

> Participant with ID, NCD-12

There was little or no experience of ‘treating to target’ i.e., reviewing the patient’s response and adapting the intervention until the target level of improvement had been achieved. No systems were available to make sure this happened. Decisions to escalate care solely relied on individual clinician assessments. The existing health information system (HMIS) does not allow tracking of treatment to target, although there was some interest in improving information use to support clinical care.

> *“There is an electronic system as a reference guideline. There are some with good experience in case of hypertension and also diabetes. So, I think it will be good if they are connected through a system and share information. I think it will be good if the system enables you to share things and information that are sometimes hard for decision making. We can deliver better service to the patient.”*

> Participant with ID, NCD-08

### Organisational readiness to implement EPHCG

All items on the ORIC received high endorsement. With regard to change-efficacy measures, ‘people who work here feel confident that the organisation can get people invested in implementing this change’ and ‘people who work here feel confident that they can manage the politics of implementing this change’ were the items with the highest (94.2%) and the lowest (89.5%) scores, respectively. For the change commitment measures, ‘people who work here are motivated to implement this change and ‘people who work here will do whatever it takes to implement this change’ were the items with the highest (96%) and the lowest (87.1%) scores respectively (Table 3).

**Table 3.** Distribution of scores of Organisational Readiness to Implement Change (ORIC)

| Characteristics | Number | Mean (standard deviation) | Response | Number (%) |
| --- | --- | --- | --- | --- |
| E1. People who work here feel confident that the organisation can get people invested in implementing this change. | 119 | 4.66 (0.82) | Disagree | 5 (4.2) |
|  |  |  | Neither agree nor disagree | 2 (1.7) |
|  |  |  | Agree | 112 (94.2) |
| C1. People who work here are committed to implementing this change.* | 108 | 4.69 (0.73) | Disagree | 3 (2.8) |
|  |  |  | Neither agree nor disagree | 2 (1.9) |
|  |  |  | Agree | 103 (95.4) |
| E2. People who work here feel confident that they can keep track of progress in implementing this change. | 121 | 4.55 (0.97) | Disagree | 8 (6.6) |
|  |  |  | Neither agree nor disagree | 2 (1.7) |
|  |  |  | Agree | 111 (91.8) |
| C2. People who work here will do whatever it takes to implement this change.* | 124 | 4.40+0.99 | Disagree | 9 (7.2) |
|  |  |  | Neither agree nor disagree | 7 (5.6) |
|  |  |  | Agree | 108 (87.1) |
| E3. People who work here feel confident that the organisation can support people as they adjust to this change. | 123 | 4.55 (0.86) | Disagree | 5 (4.1) |
|  |  |  | Neither agree nor disagree | 3 (2.4) |
|  |  |  | Agree | 115 (93.5) |
| C3. People who work here want to implement this change.* | 123 | 4.72 (0.72) | Disagree | 3 (2.4) |
|  |  |  | Neither agree nor disagree | 4 (3.3) |
|  |  |  | Agree | 116 (94.3) |
| E4. People who work here feel confident that they can keep the momentum going in implementing this change. | 119 | 4.53 (0.79) | Disagree | 4 (3.4) |
|  |  |  | Neither agree nor disagree | 4 (3.4) |
|  |  |  | Agree | 111 (93.3) |
| E5. People who work here feel confident that they can handle the challenges that might arise in implementing this change. | 124 | 4.51 (0.83) | Disagree | 4 (3.2) |
|  |  |  | Neither agree nor disagree | 6 (4.8) |
|  |  |  | Agree | 114 (91.5) |
| C4. People who work here are determined to implement this change.* | 44 | 4.59 (0.62) | Disagree | 0 (0) |
|  |  |  | Neither agree nor disagree | 3 (6.8) |
|  |  |  | Agree | 41 (93.2) |
| E6. People who work here feel confident that they can coordinate tasks so that implementation goes smoothly. | 121 | 4.51 (0.91) | Disagree | 6 (4.9) |
|  |  |  | Neither agree nor disagree | 4 (3.3) |
|  |  |  | Agree | 111 (91.8) |
| C5. People who work here are motivated to implement this change.* | 124 | 4.69 (0.68) | Disagree | 2 (1.6) |
|  |  |  | Neither agree nor disagree | 3 (2.4) |
|  |  |  | Agree | 119 (96) |
| E7. People who work here feel confident that they can manage the politics of implementing this change. | 124 | 4.44 (0.90) | Disagree | 5 (4) |
|  |  |  | Neither agree nor disagree | 8 (6.5) |
|  |  |  | Agree | 111 (89.5) |

### Relationship between ORIC and sociodemographic factors

Readiness to implement EPHCG was significantly associated with attendance at the EPHCG training (p-value 0.003) but not with any other characteristics of the health workers and their roles (Table 4).

**Table 4:** Association of ORIC score with selected background characteristics.

| Characteristics | Number (%) | Median and Interquartile range (IQR) | p-value |
| --- | --- | --- | --- |
| Gender |  |  | 0.69 |
| Male | 46 (47.9) | 53 (48,55) |  |
| Female | 50 (52.1) | 53.5 (49,55) |  |
| Age (years) |  |  | 0.98 |
| $\geq 27$ | 37 (45.1) | 53 (50,55) | |
| 20-26 | 45 (54.9) | 53 (49,55) |  |
| Profession |  |  | 0.18 |
| Health Officer/General Practitioner | 31 (32.9) | 51 (46,55) |  |
| Nurses/Midwives | 63 (67.1) | 53 (50,55) |  |
| Role at the health centre |  |  | 0.05 |
| Emergency/OPD | 47 (57.3) | 51 (46,55) |  |
| Health Centre Head | 2 (2.4) | 55 (55,55) |  |
| MCH/midwifery | 33 (40.3) | 54.5 (53,55) |  |
| Health centres by district |  |  | 0.15 |
| Sodo | 62 (64.6) | 53.5 (50,55) |  |
| Meskan/Butajira | 34 (35.4) | 51 (44,55) |  |
| EPHCG training attended |  |  | 0.003 |
| Yes | 57 (60.6) | 54 (51,55) |  |
| No | 37 (39.4) | 51 (44,55) |  |

### Synthesising barriers and enablers to implementation of EPHCG

The main contextual barriers and enablers associated with the implementation of EPHCG were mapped onto the CFIR and TDF frameworks and potential implementation strategies identified using the ERIC tool are indicated in Table 5.

**Table 5:**
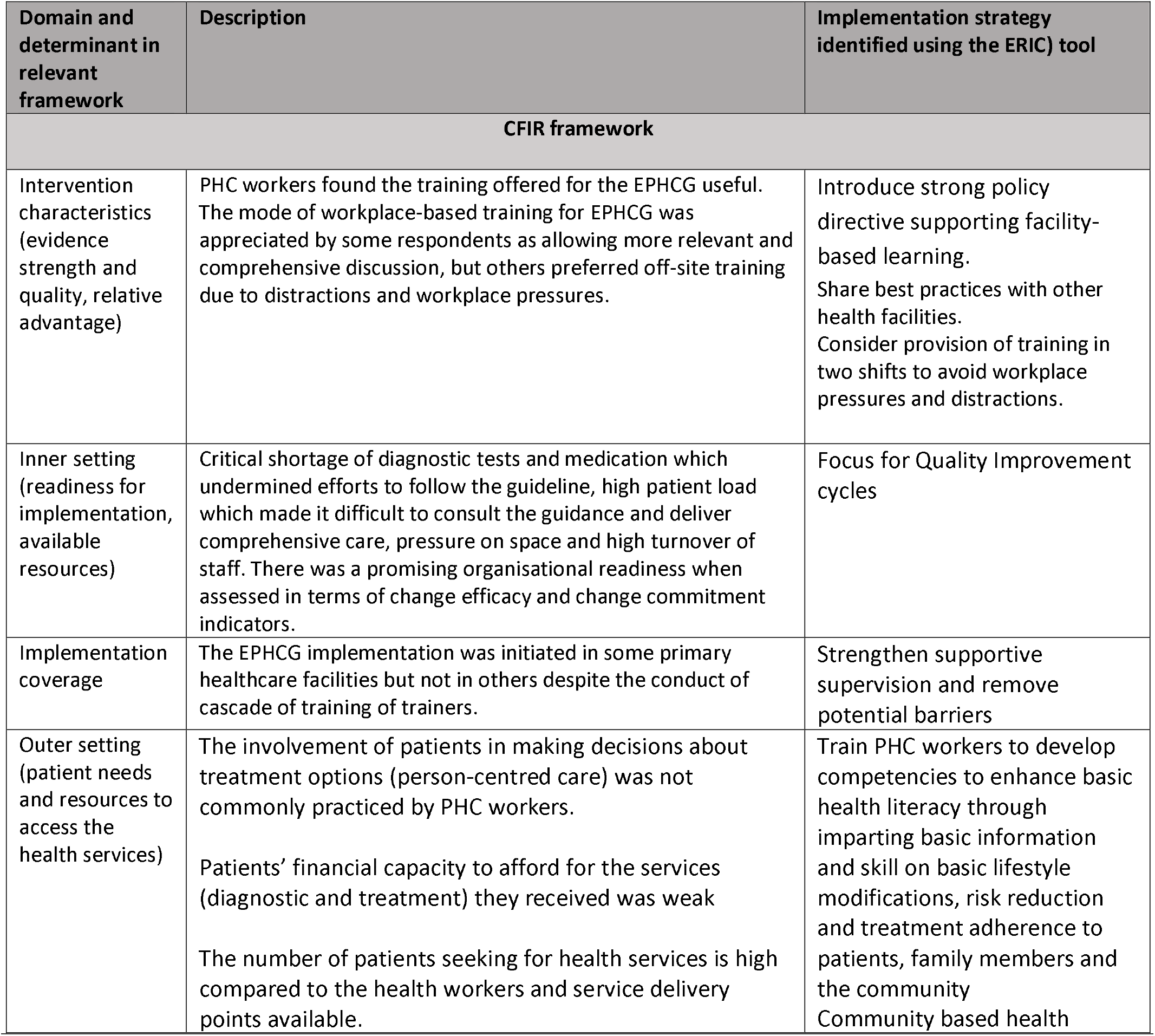
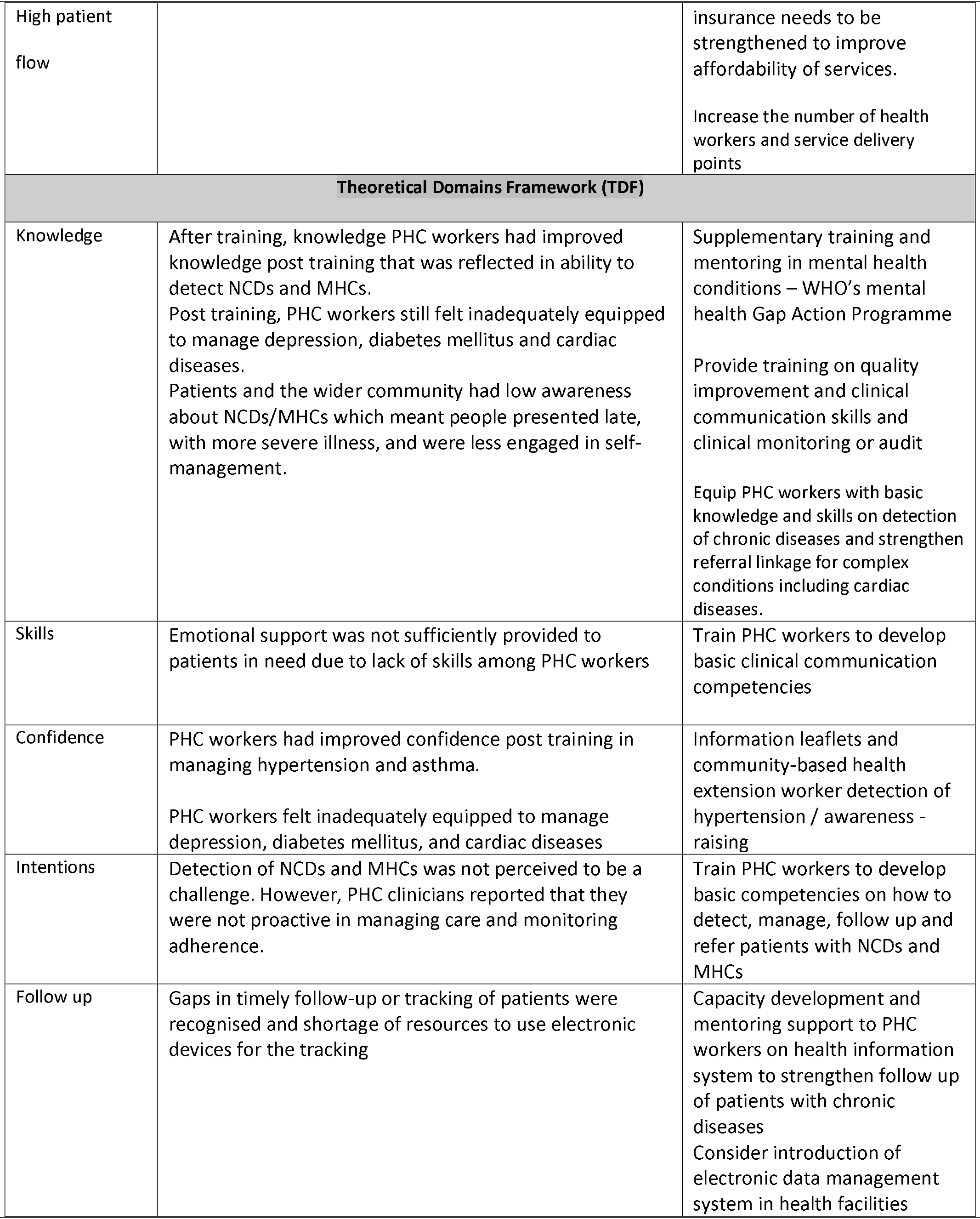
Barriers and enablers to the implementation of EPHCG identified using the Theoretical Domains Framework (TDF) and the Consolidated Framework for Implementation Research (CFIR) and implementation strategies identified using Expert Recommendations for Implementing Change (ERIC) tool

## Discussion

In this study, we investigated barriers and facilitators to the EPHCG health system strengthening initiative to improve quality of primary care and integrate care for people with NCDs/MHCs into the primary healthcare platform. Implementing EPHCG was largely seen as benefitting care quality. However, the success of its implementation varied from one health facility to another. Implementation barriers (inner setting) were largely related to facility-level resources (affecting supply) and PHC workers’ competence (impeding care pathways) and community-level awareness (affecting demand); however, organisational readiness to change was promising as a potential enabler.

Implementation of EPHCG was reported to have positively influenced healthcare delivery in PHC facilities through improving knowledge and skill in detecting and managing the care of people with NCDs and mental health conditions. This is in line with previous programmatic evaluation of EPHCG (14). However, while healthcare providers reported being confident in identifying and managing hypertension and asthma, they did not feel sufficiently competent or confident to manage depression, cardiac problems and diabetes. This is likely to be a reflection of the limited pre-service and in-service training received by primary healthcare workers for those three conditions, which have largely been managed in hospital settings prior to introduction of EPHCG. Previous experience of expanding access to mental health care in Ethiopia through integration in primary health care indicated that clinical placements as part of training, group supervision by a specialist (e.g. psychiatric nurse) and continuing quality improvement activities are needed to embed this model in the health system (33,34).

Overall, the EPHCG training model was reported to have been relevant and useful in supporting PHC workers to improve the quality of care (intervention characteristics). A similar experience has been reported from cascading of PACK training tools in PHC facilities in South Africa (35). The study participants reflected that the onsite (facility based) EPHCG training was found to engage the trainees through creating opportunities to discuss the content in a more practical and applied way (intervention characteristics). In contrast to this, the onsite training was not appreciated by some clinicians due to distraction by competing commitments and high workload. This is in line with some of the post-training evaluations of EPHCG conducted by the MoH(15).

There were challenges in training and implementing EPHCG in primary health care facilities. The principal reasons for these were lack of commitment at the health centers, lack of familiarity with peer learning, difficulty finding time to train while delivering routine clinical services, lack of financial incentives, concerns about fidelity/quality of training, turnover of trained trainers and facilitators and poor adherence in using the guidelines during consultations by the PHC workers. Inadequate technical support and supervision by the district health officers and poor awareness about EPHCG across all levels of the health system (intervention characteristics) were also barriers to successful implementation, in line with previous work (15). On the other hand, the high endorsement of indicators of organisation readiness (in terms of both change efficacy and change commitment) are potential facilitators for implementation of the changes required for EPHCG. Only attendance at EPHCG training, but no characteristics of health workers, were associated with organisational readiness. This supports the importance of the EPHCG training approach as a system strengthening intervention in its own right, bringing together facility health workers to work collaboratively in the work setting. Similar reports were made on the importance of onsite EPHCG training as documented in the post-training evaluation of EPHCG which was conducted by the health centre reform team, MoH (15).

Pertaining to awareness and patient’s needs (demand side), the barriers reported were use of traditional medicine and consequent late presentation with complications (outer setting), no support for emotional needs and disappointment with poor availability of diagnostic tests and medications (inner setting). Furthermore, low health literacy, especially in patients from rural settings was found to negatively impact implementation of person-centered care. In the post-training evaluation of EPHCG by MoH, health workers similarly reported that low community awareness undermined achieving person-centred care (15).

There were also key findings addressing the supply side. Though there is community-based health insurance system to overcome affordability and accessibility related bottlenecks in providing services, the study participants underscored these problems as a major concern to many patients with chronic diseases (outer setting). This may relate to the high out-of-pocket costs that are not covered by health insurance (e.g., for transport) and the opportunity costs accrued by repeated out-patient consultations.

Affordability of care was also identified in a study that reported the challenges and experiences of primary healthcare in 20 LMICs (36). High patient load (outer setting), shortage of space for treating and consultation, especially for mental health conditions, poor availability of laboratory tests and weak monitoring of patients’ condition, poor medication supply and shortage of health professionals were the major conditions to constrain the supply side of the EPHCG implementation as perceived by the study participants (inner setting).

In relation to the care pathways, detection of NCDs and MHCs were not perceived to be a challenge by the study participants (intentions); however, previous work in this setting has found detection to be a key bottleneck on the depression care pathway, with less than 1% of people with depression detected in primary care (37). Health workers need to promote self-management and family support to enhance lifestyle changes when counseling patients as reported by the study participants. Adherence to drugs among patients with NCDs and mental health conditions was perceived to be unsatisfactory. Follow up of patients is being addressed in some centres through enrolment into a chronic condition registry but tracing patients who disengage from care through linkages to community-based platforms such as the health extension programme is needed. Ensuring good treatment outcome or ‘treatment to target’ solely relies on individual clinicians and study participants identified that this could be strengthened through electronic data management systems. Findings in relation to indicators of care pathways showed that effective implementation of EPHCG has the potential to improve care continuum including for NCDs. All in all, the findings in all domains, i.e., EPHCG implementation, demand side, supply side and care pathways indicated that implementation of integrated primary healthcare services are still at early stages. Elsewhere, implementations of PACK, have been used across a continuum of health systems strengthening life cycles, sometimes at formative stages to test feasibility and effectiveness of decentralizing chronic disease care to primary care (38); to catalyse and create health systems demand for more comprehensive and integrated primary care (39) and in others to consolidate policy reform and aid implementation of greater task-shared care (40). Its comprehensiveness, team-based and iterative design are all intended to nurture health system strengthening efforts but must be complemented by synergistic efforts to address structural barriers such as medication and diagnostic test supply chains, need for a culture of lifelong learning among health workers and greater attention to community literacy and demand generation.

Based on the key findings thus far, we identified a range of recommendations for strengthening EPHCG implementation using the ERIC tool. These include the MoH and regional health bureaus instituting a strong policy directive supporting facility-based or on-site learning, introducing quality improvement cycles, strengthening supportive supervision and removing potential barriers in relation to medication and laboratory test supplies.

All stakeholders at different levels need to continuously train PHC workers to develop competencies (e.g., taking a blood pressure appropriately or practising good clinical communication skills). Supplementary training of EPHCG itself on MHCs (e.g. using WHO’s mental health Gap Action Programme(41) and some NCDs (diabetes and cardiac diseases) needs to be implemented by the MoH and other stakeholders. Capacity development and mentoring of PHC workers needs a special focus on person centered care and emotional support. Information leaflets and community-based health extension worker need to work on creating awareness about chronic diseases (NCDs and MHCs). Researchers need to conduct further studies with larger samples size to assess organisational readiness to implement similar public health interventions in PHC facilities.

The strengths of this study were the study design employed which enabled us to address the study objectives. Analyses of both qualitative and quantitative data were rigorously carried out. The limitations in conducting this study could be the data collection being restricted to ASSET project settings and may subsequently be difficult to transfer the findings to other primary health facilities in Ethiopia. The sample size for the quantitative study was small and there was possible social desirability bias in responding to the ORIC tool.

## Conclusion

The PHC workers reflected that EPHCG training was useful, created a sound platform for focused case-based discussion, improved knowledge and skill in detecting and managing NCDs and MHCs and was associated with higher scores of organisational readiness to change. However, the onsite training model was threatened by competing workplace pressures. Structural barriers (shortage of medications and tests, high patient caseloads, poorly developed HMIS for chronic care) and poor patient and community literacy constrained its implementation. In response, we developed additional system strengthening interventions (which could be accessed at https://cdt-africa.org/images/Downloads/Asset/On_Site_Facilitators_Guide_Training_Manual.pdf, http://repository.iifphc.org/handle/123456789/1630).

## Declaration section

### Ethical approval and consent to participate

The data collection methods were designed in line with the guidelines of research ethics principles. Ethical approval for this study was obtained from the Institutional Review Board of Addis Ababa University, College of Health Sciences, and King’s College London. Informed consent to participate in the study was obtained from all study participants.

### Availability of data and materials

All the materials are owned by the authors of this manuscript and there is none that belongs to the third party.

### Consent to publish

Not Applicable.

### Funding

The data collection underpinning the findings presented in this paper was funded by the National Institute of Health and Care Research (NIHR) Global Health Research Unit on Health System Strengthening in Sub-Saharan Africa (ASSET), King’s College London (GHRU 16/136/54) using UK aid from the UK Government. Charlotte Hanlon (C.H.) receives support through an NIHR RIGHT grant (NIHR200842) and an NIHR global health research group on homelessness and mental health in Africa (HOPE; NIHR134325). The views expressed in this publication are those of the authors and not necessarily those of the NHS, the National Institute for Health and Care Research or the Department of Health and Social Care, England. C.H. is also funded by the Wellcome Trust through grants 222154/Z20/Z (SCOPE) and 223615/Z/21/Z (PROMISE).

### Conflict of interest

None of the authors has conflict of interest.

### Authors’ contribution

A.B. wrote the main manuscript and did both qualitative and quantitative analyses. A.A. reviewed the manuscript. N.Sew. reviewed the manuscript.

T.E. facilitated the data collection and organised the data set. T.G. reviewed the manuscript. Y.G. reviewed the manuscript. W.M. reviewed the manuscript. G.M reviewed the manuscript. L.F. reviewed the manuscript. N.Sev. reviewed the manuscript. M.P. reviewed the manuscript. A.F. reviewed the manuscript. C.H. reviewed the manuscript and supported the data management and analyses

## Data Availability

The data for this manuscript is available upon reasonable request.

## Acknowledgment

The authors would like to acknowledge NIHR, King’s College London, Wellcome Trust for funding the study. We also would like to appreciate C.H. for her outstanding contribution in facilitating the fund request and realising the conduct of the study. The qualitative study findings of this study on implementation of primary healthcare clinical guidelines (Oral-Poster) were presented at the scientific conference of implementation science which was organized by the King’s College in London.

## Notes

### Competing Interest Statement

The authors have declared no competing interest.

### Funding Statement

National Institute for Health and Care Research, King's College London and Wellcome Trust.

### Author Declarations

Addis Ababa University, College of Health Sciences Institutional Review Board

